# A Machine Learning Model Incorporating Laboratory Blood Tests Discriminates Between SARS-CoV-2 and Influenza Infections at Emergency Department Visit

**DOI:** 10.1101/2021.08.06.21261713

**Authors:** Junyi Liu, Lars F. Westblade, Amy Chadburn, Richard Fideli, Arryn Craney, Sophie Rand, Melissa Cushing, Zhen Zhao, Jingjing Meng, He S. Yang

**Author notes:** Joint corresponding authors <u>Correspondence to:</u> He S. Yang, PhD, Department of Pathology and Laboratory Medicine, Weill Cornell Medicine, 525 E. 68th Street, Suite F 707, New York, NY 10065, Jingjing Meng, PhD, Department of Computer Science and Engineering, University of Buffalo, 304 Davis Hall, Buffalo. NY.

## Abstract

**Introduction:** Severe acute respiratory syndrome coronavirus 2 (SARS-CoV-2) and influenza virus are contagious respiratory pathogens with similar symptoms but require different treatment and management strategies. This study investigated whether laboratory blood tests can discriminate between SARS-CoV-2 and influenza infections at emergency department (ED) presentation.

**Methods:** 723 influenza A/B positive (2018/1/1 to 2020/3/15) and 1,281 SARS-CoV-2 positive (2020/3/11 to 2020/6/30) ED patients were retrospectively analyzed. Laboratory test results completed within 48 hours prior to reporting of virus RT-PCR results, as well as patient demographics were included to train and validate a random forest (RF) model. The dataset was randomly divided into training (2/3) and testing (1/3) sets with the same SARS-CoV-2/influenza ratio. The Shapley Additive Explanations technique was employed to visualize the impact of each laboratory test on the differentiation.

**Results:** The RF model incorporating results from 15 laboratory tests and demographic characteristics discriminated SARS-CoV-2 and influenza infections, with an area under the ROC curve value 0.90 in the independent testing set. The overall agreement with the RT-PCR results was 83% (95% CI: 80-86%). The test with the greatest impact on the differentiation was serum total calcium level. Further, the model achieved an AUC of 0.82 in a new dataset including 519 SARS-CoV-2 ED patients (2020/12/1 to 2021/2/28) and the previous 723 influenza positive patients. Serum calcium level remained the most impactful feature on the differentiation.

**Conclusion:** We identified characteristic laboratory test profiles differentiating SARS-CoV-2 and influenza infections, which may be useful for the preparedness of overlapping COVID-19 resurgence and future seasonal influenza.

## Introduction

Both the severe acute respiratory syndrome coronavirus 2 (SARS-CoV-2), the etiology agent of coronavirus disease 2019 (COVID-19), and influenza virus are contagious respiratory pathogens, which cause as a wide range of illnesses from asymptomatic or mild through to severe disease and death.^1^ However, the fraction of severe infection cases, as well as the mortality rate, is higher in COVID-19 than in influenza.^2^ Transmissibility, estimated by the basic reproductive rate (R_2_), is also higher in SARS-CoV-2 (R_0_ ~2.5) compared to influenza (R_0_ ~1.7).^3^ The SARS-CoV-2 and influenza virus infections have similar symptoms, such as cough, sore throat, fever, fatigue, and myalgias, making clinical differentiation at hospital presentation challenging without the aid of laboratory tests. Currently, the detection of these pathogens relies on viral specific real-time reverse-transcription polymerase chain reaction (RT-PCR) testing of nasopharyngeal swabs or other upper respiratory track specimens.^4^ However, while the turn-around time (TAT) of RT-PCR testing is usually within a day, it can be substantially longer due to reagent shortages^5^ as experienced during the COVID-19 pandemic, or lack of on-site testing, particularly in many of the smaller and more rural hospitals. Since the treatment of SARS-CoV-2 and influenza is different, delayed identification of infected patients may result in inappropriate patient management and increased risk of infection for healthcare personnel. Therefore, rapid discrimination of SARS-CoV-2 and influenza infections, and identification of high-risk patients is vital for individual patient care and for controlling disease transmission.

Routine laboratory tests provide objective and quantifiable characterization of the effects of the virus on the human body^6^. Routine test results are generally available within 1-2 hours and are accessible prior to patient discharge from the emergency department (ED). Previous studies^7, 8^, including our own publications^9^, have demonstrated that machine learning models incorporating routine laboratory blood tests and demographics can differentiate and predict SARS-CoV-2 infected and non-infected patients. Machine learning algorithms have the capability of revealing complicated pattern or trend behind high-dimensional laboratory data that are challenging for human eyes to visualize. So far, limited studies have investigated the differences in laboratory blood test results between SARS-CoV-2 and influenza infections^3, 10^. While no single laboratory test strongly associates with SARS-CoV-2 or influenza infection, we investigated whether a combination of multiple routine laboratory tests can discriminate between these two viral infections at an early stage of disease, and aimed to find out the most impactful laboratory features that contribute to the discrimination. Furthermore, COVID-19 has been evolving since the initial outbreak of the disease^11^. We are interested in understanding whether the impactful laboratory features remain unchanged in more recent COVID-19 positive patients during the 2020-2021 winter wave.

## Methods

### Patient cohorts

We reviewed retrospectively laboratory and demographic data of 723 influenza A/B positive patients from January 1, 2018 to March 15, 2020, and 1,281 SARS-CoV-2 positive patients from March 11, 2020 to June 30, 2020, evaluated in the ED of a New York City academic hospital. Routine influenza RT-PCR testing was suspended from March to September 2020 to prioritize resources for SARS-CoV-2 testing. The SARS-CoV-2 cohort included infected patients from the initial outbreak as well as from the post-apex phase. Influenza or SARS-CoV-2 RT-PCR results, routine laboratory testing results within 2 days prior to the completion of RT-PCR testing, and patient demographic information were obtained from the laboratory information system (Cerner Millennium, Cerner Corporation). Exclusion criteria included patients < 18 years of age, and patients who lacked laboratory test results within the designated time window (within 2 days prior to RT-PCR results release). In total, the final dataset included results from 1,237 SARS-CoV-2 RT-PCR positive patients and 513 influenza positive (393 type A, and 119 type B, 1 A/B) ED patients (**Figure 1**). This study was approved by the Institutional Review Board (IRB) of Weill Cornell Medicine and deemed IRB exempt by the University of Buffalo.

**Figure 1:**
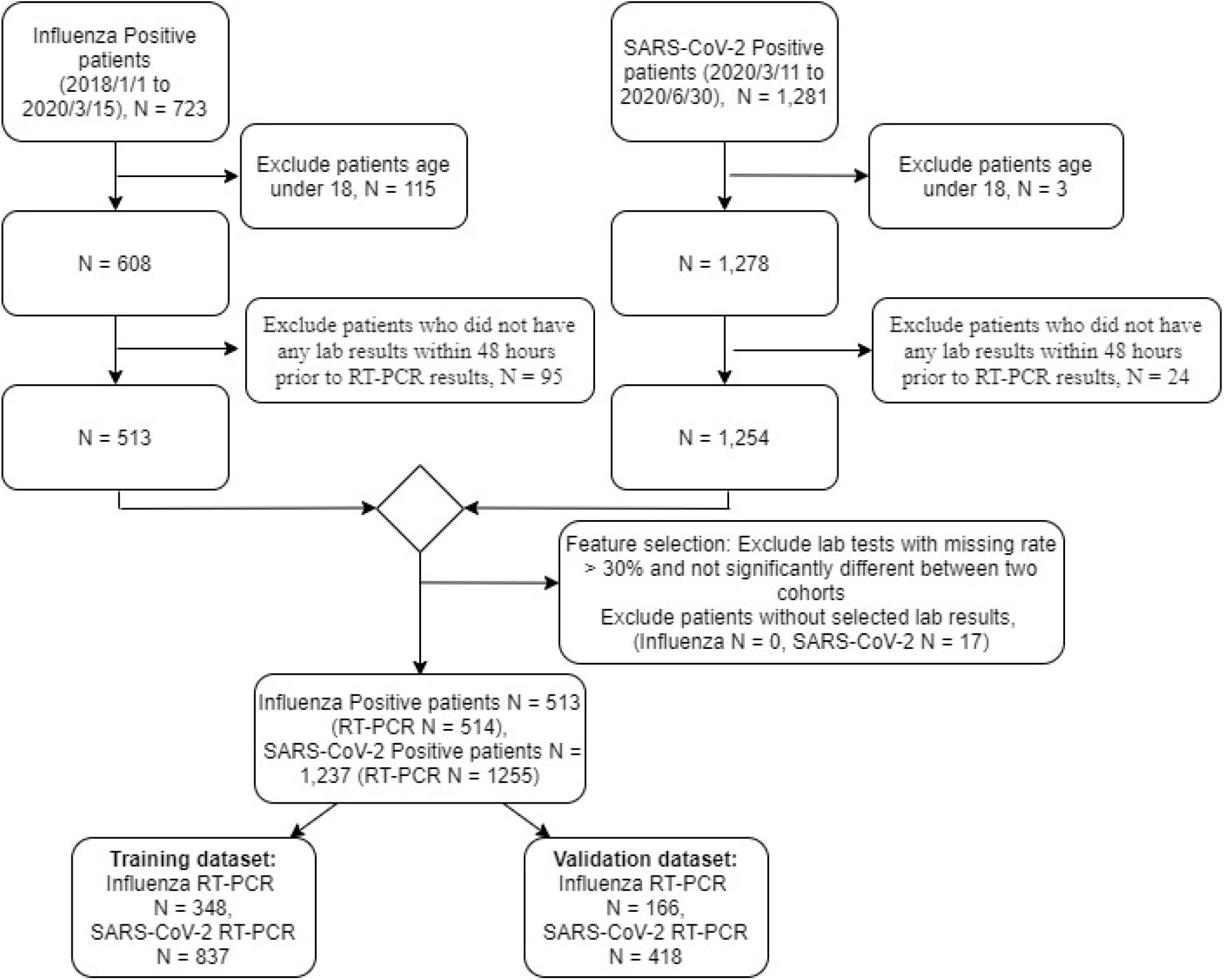
Inclusion/exclusion cascade of SARS-CoV-2 and influenza patient cohorts in the dataset.

From December 1, 2020 to February 28, 2021, there were a total of 559 SARS-CoV-2 RT-PCR positive patients and only 1 influenza RT-PCR positive patient (this patient was also positive for SARS-CoV-2) who were evaluated in the ED of our hospital. After applying the exclusion criteria as state above, 519 SARS-CoV-2 positive patients were included in the new dataset. Since there were only 1 positive influenza case during the same time, we had to compare the new SARS-CoV-2 dataset to the previous influenza dataset from 2018/1/1 to 2020/3/11.

### Model construction

A total of 57 chemistry, hematology, and coagulation tests commonly ordered in these patients were examined in the SARS-CoV-2 and influenza cohorts. Of them, 15 laboratory tests were selected to be included in the model based on two criteria: 1) a result available for at least 70% of the patients within 48 hours before a specific SARS-CoV-2 RT-PCR test and 2) showing a significant difference (P-value after Bonferroni correction less than 0.05) between influenza and SARS-CoV-2 positive patients. After the feature selection, a 22-dimensional vector (15 laboratory tests, one age, one gender, five race variables (African American, Asian, Caucasian, others and unknown) was constructed to represent every SARS-CoV-2 or influenza RT-PCR test result (**Figure 2**). If one laboratory test was ordered multiple times within 48 hours, an average of the values was calculated and used for analysis. The missing value of a specific laboratory test in a feature vector was imputed by the mean value of the available non-missing values of that dimension over all patients. Subsequently, a random forest classifier model was developed incorporating the results of 15 selected laboratory tests and patient age, gender, and race, using the Python scikit-learn package 0.23.2. The whole data set was randomly split into a training set (2/3 of cases) and a testing set (1/3 cases) with the same ratio of SARS-CoV-2/influenza cases as the ratio for the overall cases. The hyperparameters of random forest model was trained using 5-fold cross validation, where all RT-PCR tests in the training set were randomly partitioned into 5 equal buckets with the same SARS-CoV-2/influenza ratio. The random forest model was then trained using selected hyperparameters with all RT-PCR tests in the training set. The performance of the model was evaluated in the independent testing set using area under the receiver operating characteristic curve (ROC), sensitivity, specificity, and agreement with RT-PCR. The Shapley Additive Explanations (SHAP) technique was employed to visualize the impact of each laboratory test on the differentiation.

**Figure 2:**
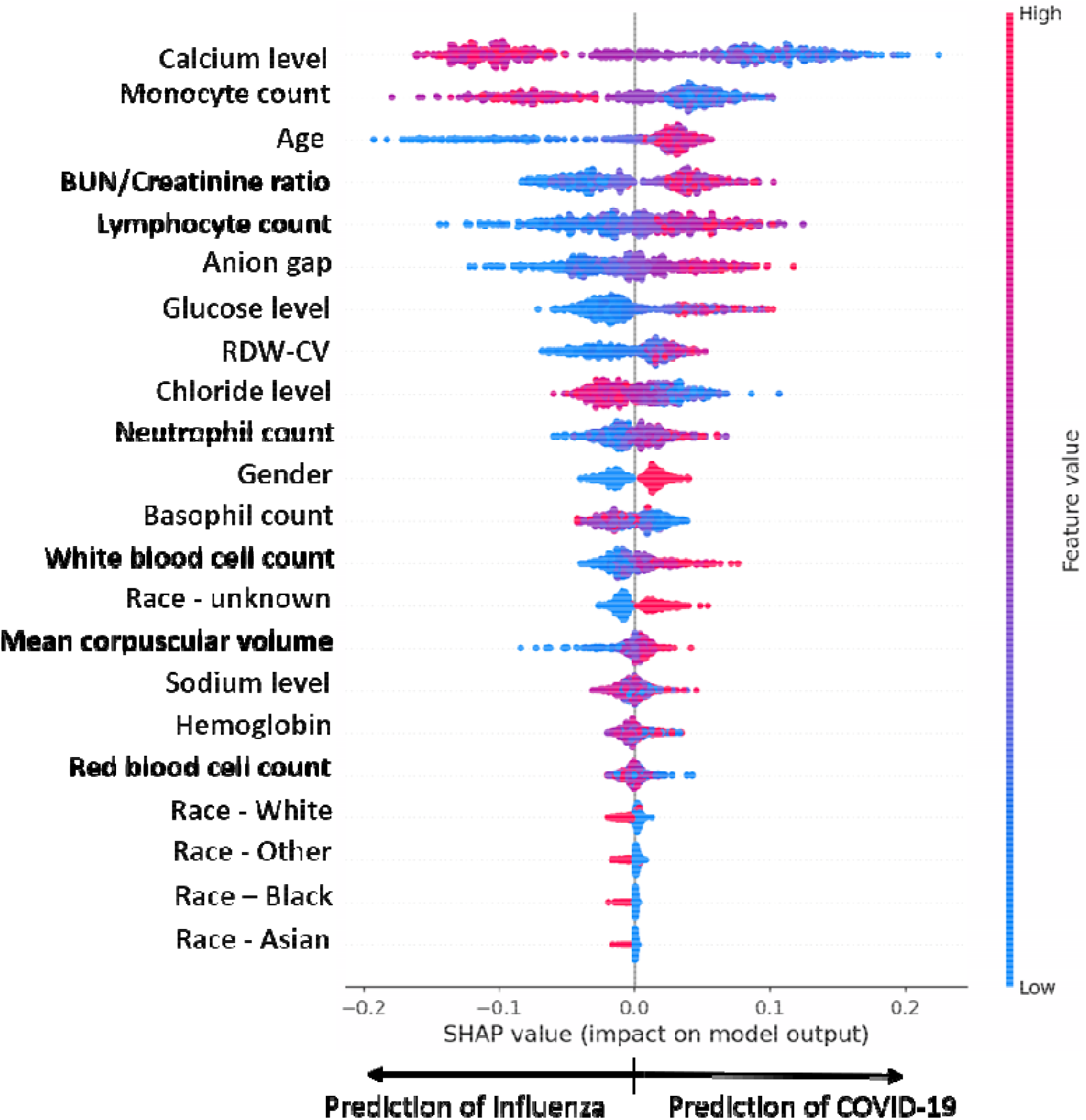
Impact of each laboratory test and demographic feature to the overall differentiation of SARS-CoV-2 and influenza infections in the random forest model. Dataset includes SARS-CoV-2 positive patients from 2020/3/15 to 2020/6/30, and influenza positive patients from 2018/1/1 to 2020/3/11. Laboratory tests are organized on the y-axis according to their impact from high to low. Individual values of each test for each patient are colored according to their relative values. Higher and lower values are shown in red and blue, respectively. Absolute SHAP values of each test are shown on the x-axis. Positive SHAP value to the right indicates prediction of SARS-CoV-2 whereas negative SHAP value to the left indicates prediction of influenza.

## Results

Patients with SARS-CoV-2 infection were significantly older (median: 64 years, IQR: 51-75) than those with influenza (median: 48 years, IQR: 32-63, p < 0.001). The percentage of male patients was also significantly higher in SARS-CoV-2 cohort (58.36%) than in the influenza group (41.33%, p < 0.001). In addition, a significantly lower percentage of African American was present in the SARS-CoV-2 group than the influenza cohort (p = 0.02, **Table 1**).

**Table 1.** Demographic of SARS-CoV-2 and influenza patient cohorts

| Demographics: | SARS-CoV-2 positive patients from 2020/3/11 to 2020/6/30 (n = 1,237) | Influenza positive patients from 2018/1/1 to 2020/3/15 (n = 513) | SARS-CoV-2 positive patients from 2020/12/1 to 2021/2/28 (n = 519) |
| --- | --- | --- | --- |
| Age, years; median (IQR) | 64 (51-75) | 48 (32-63)<sup>*#</sup> | 61 (45-73) |
| Gender, male, n (%) | 722 (58.36%) | 212 (41.33%)<sup>*#</sup> | 253 (48.75%) |
| Race: |  |  |  |
| Asian | 52 (4.20%) | 32 (6.24%) | 33 (6.36%) |
| African American | 117 (9.46%) | 66 (12.87%)<sup>*</sup> | 71 (13.68%) |
| White | 324 (26.19%) | 153 (29.82%)<sup>#</sup> | 21 (40.66%) |
| Other | 213 (17.22%) | 128 (24.95%)<sup>*#</sup> | 93 (17.92%) |
| Unknown | 531 (42.93%) | 134 (26.12%)<sup>*#</sup> | 111 (21.39%) |

<sup>\*</sup>p value < 0.05 between SARS-CoV-2 positive patients from 2020/3/11 to 2020/6/30 and influenza positive patients from 2018/1/1 to 2020/3/15.
<sup>#</sup>p value < 0.05 between SARS-CoV-2 positive patients from 2020/12/1 to 2021/2/28 and influenza positive patients from 2018/1/1 to 2020/3/15.

The laboratory results completed within 48 hours prior to a positive SARS-CoV-2 or influenza RT-PCR test result were included in the analysis. A total of 15 laboratory test results with missing rate < 30% were found to be significantly different between the SARS-CoV-2 and influenza infected patients at their ED presentation. The performance of the random forest model incorporating these 15 laboratory tests and patient age, sex and race achieved an area under the ROC curve (AUC) value of 0.90 (95% CI: 0.88-0.93) in the independent testing set. The overall agreement with the RT-PCR, determined at the Youden’s index point, was 83% (95% CI: 80-86%), with 87% (95% CI: 84-90%) agreement with SARS-CoV-2 RT-PCR results and 73% (95% CI: 66-80%) agreement with the influenza RT-PCR results. The random forest model generated a probability score, from 0 to 1, to differentiate influenza vs. SARS-CoV-2 infection (cutoff 0.58 at the Youden’ index point).

In order to visualize and interpret the impact of each laboratory test to the overall differentiation, a Shapley Additive Explanations (SHAP) technique (Version 4.10.0) was employed which assigns each feature a value of importance (the SHAP value) for the differentiation of SARS-CoV-2 or influenza infections. A summary plot is shown in **Figure 2**, in which the impact of each laboratory test and demographic feature was ordered from high to low. Notably, serum calcium level had the greatest impact on the differentiation of these two infections. The SARS-CoV-2 patients demonstrated significantly lower serum calcium concentrations than the influenza positive patients. In addition, the peripheral monocyte and basophil counts were significantly lower whereas the lymphocyte and white blood cell counts were significantly higher in the SARS-CoV-2 patients compared to the influenza patients. The SARS-CoV-2 patients also demonstrated hypochloremia, hyperglycemia, higher BUN/creatinine ratio and higher anion gap compared to the influenza patients.

Since the COVID-19 has been evolving since the initial outbreak, we further investigated whether our random forest model is still discriminative between SARS-COV-2 and influenza in the 2020-2021 winter wave and whether the top laboratory tests are still impactful on the differentiation. From December 1, 2020 to February 28, 2021, there were a total of 559 SARS-CoV-2 RT-PCR positive patients evaluated in the ED of our hospital, and among them, 519 were included in our dataset after the exclusion criteria was applied (Demographic information in **Table 1**). However, there was only one positive influenza case identified in the ED during this time period, and this patient was also positive for SARS-CoV-2 RT-PCR testing. Therefore, we test our model on the new SARS-CoV-2 dataset with the previous 723 influenza A/B positive patients from January 1, 2018 to March 15, 2020. The random forest model with the same parameters achieved an AUC of 0.82 with an overall agreement of 74% (95% CI: 71-77%) using the same cutoff value. Agreement with SARS-CoV-2 and influenza RT-PCR results was 75% and 73%, respectively. Interestingly, serum calcium level remains the most impactful laboratory feature on the differentiation of SARS-CoV-2 and influenza infections (**Figure 3**). Compared to the influenza positive patients, the more recent SARS-CoV-2 positive patients still exhibited hypocalcemia, lower level of monocyte counts, and higher levels of lymphocyte, WBC, and neutrophil counts as well as higher anion gap and BUN/creatinine ratio.

**Figure 3:**
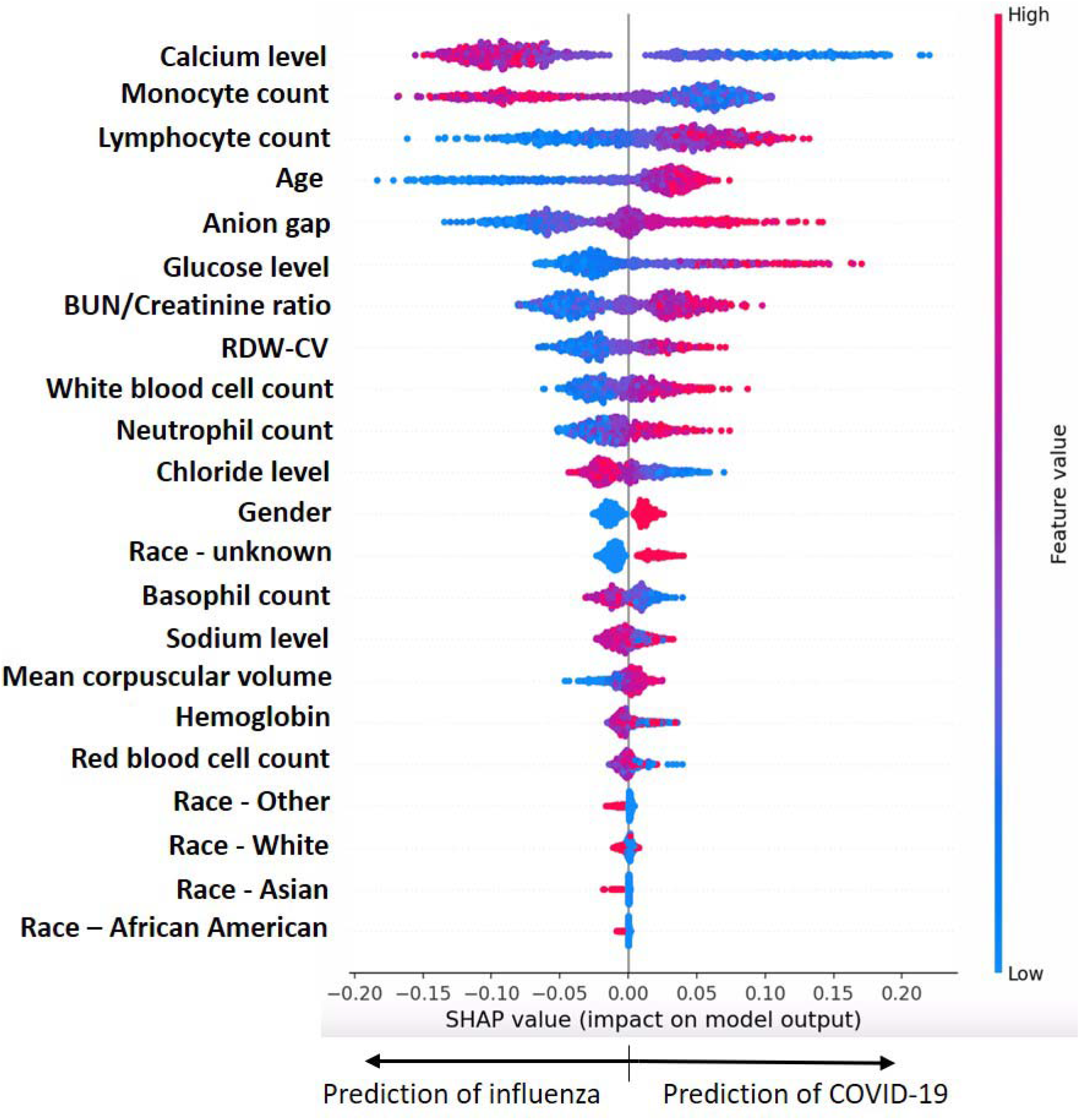
Impact of each laboratory test and demographic feature to the overall differentiation in the new SARS-CoV-2 dataset from 2020/12/1 to 2021/2/28 and the previous influenza dataset from 2018/1/1 to 2020/3/11.

## Discussion

This retrospective analysis illustrates that a combination of 15 laboratory tests, along with patient demographics, can help distinguish between SARS-CoV-2 and influenza infections at an early stage of disease. While SARS-CoV-2 and influenza infections show similar symptoms^2^, each has a characteristic laboratory test result profile that could be used for early disease differentiation and prediction. Although the level of seasonal influenza was extremely low in the U.S. and other countries in the northern hemisphere during the past winter^12, 13^, our findings pave the way for future research about the distinct pathophysiological mechanisms underlying SARS-CoV-2 and other respiratory virus infections, and may be useful for the preparedness of overlapping COVID-19 resurgence and future seasonal influenza. Understanding the characteristic laboratory profiles associated with the two pathogens may assist early and rapid identification of high-risk patients and optimize resource use during periods of high infection rates for both SARS-CoV-2 and influenza, especially in areas where hospitals or clinics do not have onsite RT-PCR testing for respiratory viruses.

Machine learning models can perform analysis of massive quantities of data and extract hidden patterns that that would be challenging for human eyes to identify. A machine learning classification model should make clinical and biologic sense. Here we report that in our machine learning classification model, the serum total calcium level, among all routine laboratory tests, has the highest impact on the differentiation of SARS-CoV-2 from influenza infections. Hypocalcemia has been previously reported as a prevalent biochemical abnormality in SARS-CoV-2 infected patients with a marked negative influence on disease severity, biochemical inflammation and thrombotic markers^14^, and has been proposed as an independent risk factor for hospitalization due to COVID-19^15^. While the exact underlying biologic mechanism for hypocalcemia in SARS-CoV-2 infection is still unclear, it may be closely associated with viral-associated multi-organ damage and increase in inflammatory cytokines^16^. Studies better delineating the mechanism and impact of viral infections on calcium metabolism may suggest therapeutic modalities in the future. In addition, SARS-CoV-2 patients exhibited lower monocyte and basophil counts, higher white blood cell, neutrophil and lymphocyte counts, as well as hypochloremia, hyperglycemia, higher BUN/creatinine ratio and higher anion gap, compared to influenza positive patients. The substantial differences in laboratory result profile between SARS-CoV-2 and influenza patients shed new lights on the reactions of human body to the two types of viral infections, which may contribute to the different disease manifestations and outcomes.

The COVID-19 winter wave in New York was not as severe as the devastating first wave that hit the city in March and April 2020. The number of hospitalized patients as well as the mortality rates were lower than those of the first wave^17^. More patients showed mild or asymptomatic disease, which may partly explain why it was more difficult to differentiate SARS-CoV-2 and influenza infections from laboratory test results and why the performance of our model with the same parameters has dropped in the new dataset. The performance of the model can be improved with a continuous learning process that involves model updating and parameter optimizing using the new data. Interestingly, despite the evolvement of COVID-19, the laboratory features that have greatest impact on the differentiation between the two viral infections have been stable in the longitudinal datasets.

A study limitation is that our model’s performance has not been validated in a dataset including concurrent SARS-CoV-2 and influenza positive patients as Influenza RT-PCR testing was suspended from March to September 2020 to prioritize resources for SARS-CoV-2 testing. We attempted to collect new data from November 2020 to February 2021, however, there was only one influenza positive case during this time in our hospital ED. This observation was consistent with the extremely low level of seasonal influenza in North America^12^. Despite a lack of direct comparison, the characteristic profile of SARS-CoV-2 in comparison to influenza infection is still valid and has the potential to impact patient care. The performance of our model could be further improved when it is trained with more concurrent influenza and SARS-CoV-2 patient data.

## Conclusion

The proposed random forest model incorporating age, gender, race and 15 routine laboratory tests can discriminate between SARS-CoV-2 and influenza infection during an initial patient ED visit. We identified 15 routine laboratory blood tests, which assist in separating of SARS-CoV-2 from influenza infection, with serum calcium level being the most impactful feature. These characteristic laboratory test result profiles associated with SARS-CoV-2 and influenza infections mirror the biologic effects of these viruses on patients and provides clinically an opportunity for early identification of high-risk patients. Our analysis demonstrates the utility of machine learning as an emerging technique to support the diagnosis of infectious diseases.

## Data Availability

The data are not available due to patient ethical restrictions.

## Abbreviation

AGAP: anion gap
COVID-19: corona virus disease-2019
SARS-CoV-2: severe acute respiratory syndrome coronavirus 2
TAT: turn-around time
ED: emergency department
RT-PCR: real-time reverse transcription polymerase chain reaction
WBC: white blood cells
RBC: red blood cells
RDW-CV: Red blood cell distribution width
BUN: Blood urea nitrogen
MCV: Mean corpuscular volume
AUC: Area under the receiver operating characteristic curve
NYPH/WCM: New York Presbyterian Hospital/Weill Cornell Medicine
NYPH/LMH: New York Presbyterian Hospital/Lower Manhattan Hospital

## Author contribution

JL for performing data analysis and manuscript editing; LFW for providing influenza data and manuscript editing; AC for manuscript editing; RF for organizing laboratory test results for COVID-19 and influenza patients; AC for manuscript editing; SR for providing COVID-19 data; MC and ZZ for manuscript editing; JM for supervision of data analysis and manuscript editing; HSY for conceptualization, project supervision, data analysis, and writing and editing of the manuscript.

